# Droplet Digital PCR as a First-Line Detection Tool in the Genetic Diagnosis of Vascular Anomalies

**DOI:** 10.64898/2026.08.11.26359368

**Authors:** Tara K. Lane, Timothy E. Green, Denisse Garza, Natasha J. Brown, Michelle G. de Silva, Mark F. Bennett, Caitlin Tubb, Sian M.W. Macdonald, Adam Gascoigne, Roderic J. Phillips, John Slavin, Colleen D’Arcy, Duncan MacGregor, Amelia Clifford, Lydia Pathmanathan, Susan J. Robertson, Phillip Bekhor, Jodie Simpson, Samuel Gooley, Ingrid E. Scheffer, Samuel F. Berkovic, Anthony J. Penington, Michael S. Hildebrand

## Abstract

Targeted precision therapies are increasingly used in the treatment of individuals with vascular anomalies (VAs). This increases the need for rapid, accurate and inexpensive genetic diagnosis. Droplet digital polymerase chain reaction (ddPCR) is an alternative to next-generation sequencing (NGS), permitting rapid, highly sensitive interrogation of recurrent pathogenic mosaic variants. We examined the feasibility of ddPCR as a primary diagnostic tool in a large cohort of individuals with VAs. Lesional tissue was collected for ddPCR of up to 46 recurrent pathogenic variants across 16 genes associated with VAs. Specimens were assessed on a subset of assays for each individual based on clinical phenotype. Most individuals who had negative ddPCR results went on to high-depth gene panel or deep exome NGS, or Sanger sequencing. Here we report the phenotypic and molecular findings for 78 newly recruited and tested individuals in addition to the 60 individuals already reported from our cohort. The overall diagnostic yield for our cohort when combined with individuals previously reported was 104/138 (75%). Of 138 individuals tested, recurrent pathogenic variants were detected in 71 (51%) on ddPCR. Variants were most frequently identified in *PIK3CA* (n=28), *TEK* (n=18), *GNAQ* (n=12), or *MAP2K1* (n=7). In a further 33 individuals, pathogenic variants were identified on NGS or Sanger sequencing. Our findings indicate that ddPCR is an efficient method achieving a high diagnostic yield in our cohort when used prior to sequencing.

## Introduction

Vascular anomalies (VA) are a group of congenital conditions affecting the development of blood and lymphatic vessels.^1^ They are broadly classified as highly-proliferative vascular tumours or slow-growing vascular malformations. The latter group comprise slow-flow capillary malformations (CMs), venous malformations (VMs) and lymphatic malformations (LMs); or fast-flow arteriovenous malformations (AVMs).^1^ Slow-flow vascular malformations are typically associated with pathogenic variants in the PI3K/AKT/mTOR pathway, while fast-flow vascular malformations are usually associated with pathogenic variants in the Ras-MAPK pathway.^1–3^ Most individuals have a non-syndromic lesion. Some individuals have syndromic forms including in the *PIK3CA*-related overgrowth spectrum (PROS) encompassing numerous phenotypes that manifest as a consequence of somatic mosaic variants in *PIK3CA*. Individuals with PROS may have features that overlap with those in the VA spectrum including the presence of slow-flow malformations and overgrowth.^1,4^ PTEN hamartoma tumour syndrome (PHTS) is a multi-system disorder driven by loss-of-function pathogenic variants in the *PTEN* tumour suppressor gene. Slow- or fast-flow vascular anomalies are present in a subset of individuals with PHTS.

Vascular anomalies are phenotypically heterogeneous, ranging in presentation and severity from simple port-wine stains associated primarily with cosmetic concerns to complex life-threatening lesions at risk of causing airway impaction or haemorrhage.^2,3^ Individuals frequently require complex medical management involving a range of healthcare specialties, and many do not incorporate pathways for genetic testing.^5–7^ Although this is slowly changing as recognition of the genetic basis of these disorders improves, limited access to genetic testing makes uncovering the genetic architecture challenging.^5,8^ Further complicating testing is the complex genetic architecture of vascular lesions. While some individuals have predisposing germline pathogenic variants in every cell, most individuals (> 80%) have lesions arising from somatic mosaicism, with causative variants restricted to a subset of cells within the lesion.^8^ Pathogenic somatic variants can be present at very low variant allele fraction (VAF; i.e., < 1%)^9–11^, requiring highly sensitive genetic testing of resected or biopsied lesional tissue for detection.^1^ Thus testing is limited by the availability of lesional tissue and access to sensitive testing technologies such as droplet digital polymerase chain reaction (ddPCR) or high-depth next generation sequencing (NGS).^1,10^

Establishing a molecular diagnosis is critical for optimal clinical management of individuals with VAs because new and repurposed targeted therapies are increasingly becoming available.^12^ Gene pathway-specific therapies, such as the mTOR inhibitor sirolimus or PI3K inhibitor alpelisib, are being used to target the PI3K/AKT/mTOR pathway in individuals with slow-flow lesions.^13^ MEK inhibitors, targeting the Ras/Raf/MAPK pathway, are being trialled in individuals with fast-flow lesions including trametinib^14^, mirdametinib (NCT05983159)^15^, and cobimetinib (NCT05125471).^16^

Here we demonstrate the diagnostic utility of ddPCR as a first-line genetic test in a large cohort of individuals with VAs including clinically complex or atypical cases. Rapid and accurate molecular diagnosis is critical to increasing access to appropriate targeted therapies.

## Results

We studied 138 individuals with VAs comprising 104 with slow-flow malformations (∼75%), 19 with fast-flow malformations (∼14%), 10 with mixed fast-flow and slow-flow lesions (∼7%), and 5 with vascular tumours (∼4%). Of these, 9 (∼7%) had PROS (Table 1). Surprisingly, six individuals (∼4%) had PHTS confirmed by discovery of a *PTEN* pathogenic variant including one with mosaicism (Table 1). We previously performed genetic analysis on 60 of these individuals and reported pathogenic variants in 33 (∼55%).^1^ Here we report updated molecular testing for 105 unsolved individuals including 27 individuals lacking a pathogenic finding from our previous study,^1^ as well as 78 individuals newly recruited to our cohort (Figure 1).

**Figure 1.**
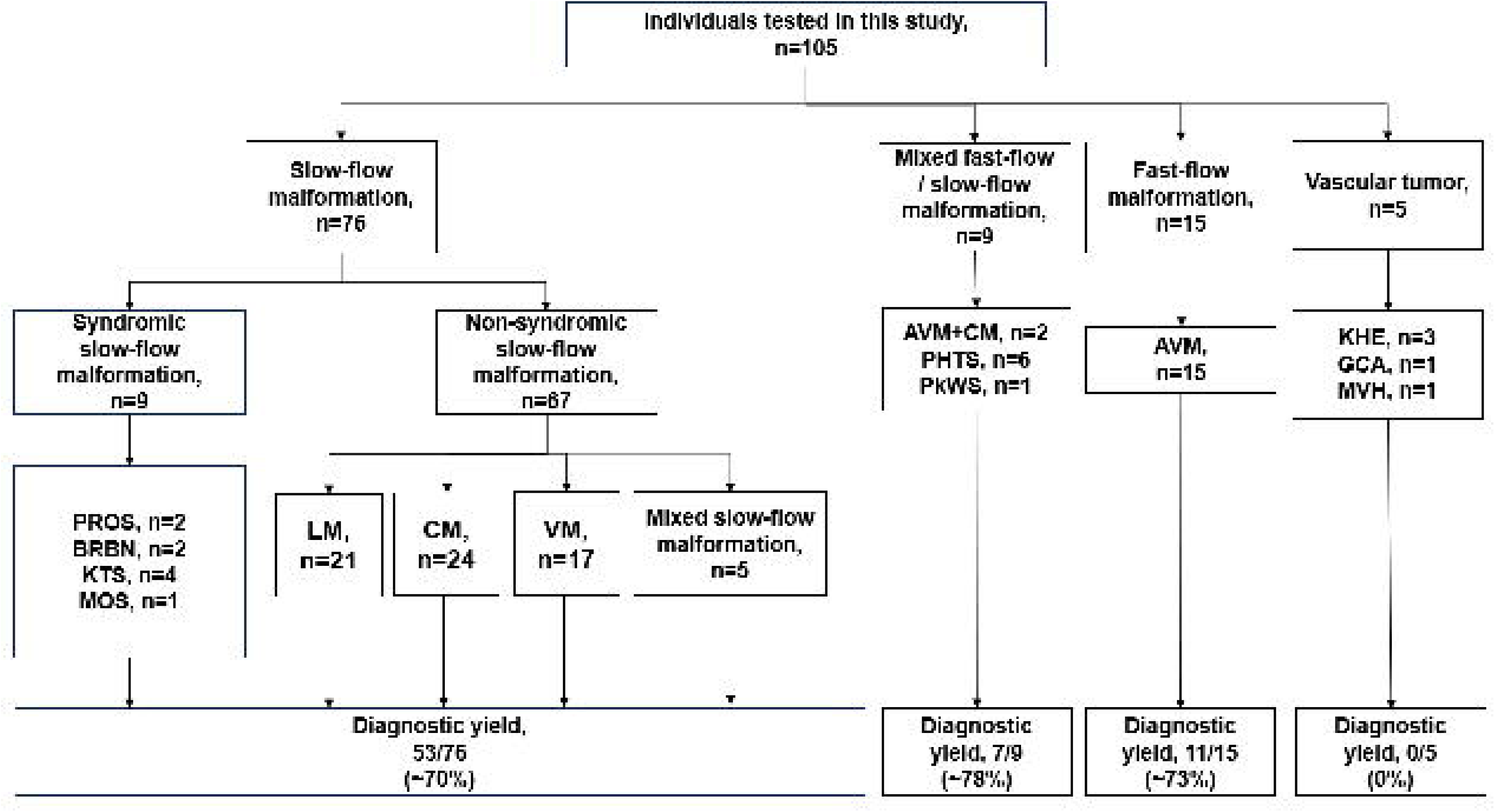
Cohort of individuals with vascular anomalies or PROS. Flow chart showing phenotypes of 105 individuals including 78 new cases and 27 unsolved cases from our previous study.^1^ Breakdown of diagnostic yield by malformation type is also shown. AVM, arteriovenous malformation; BRBN, blue rubber bleb nevus syndrome; CM, capillary malformation; GCA, giant cell angioblastoma; KHE, kaposiform haemangioendothelioma; KTS, Klippel-Trenaunay syndrome; LM, lymphatic malformation; MOS, mosaic overgrowth syndrome; MVH, microvenular haemangioma; PHTS, PTEN hamartoma tumour syndrome; PkWS, Parkes-Weber Syndrome; PROS, PIK3CA-related overgrowth spectrum; VM, venous malformation.

**Table 1.**
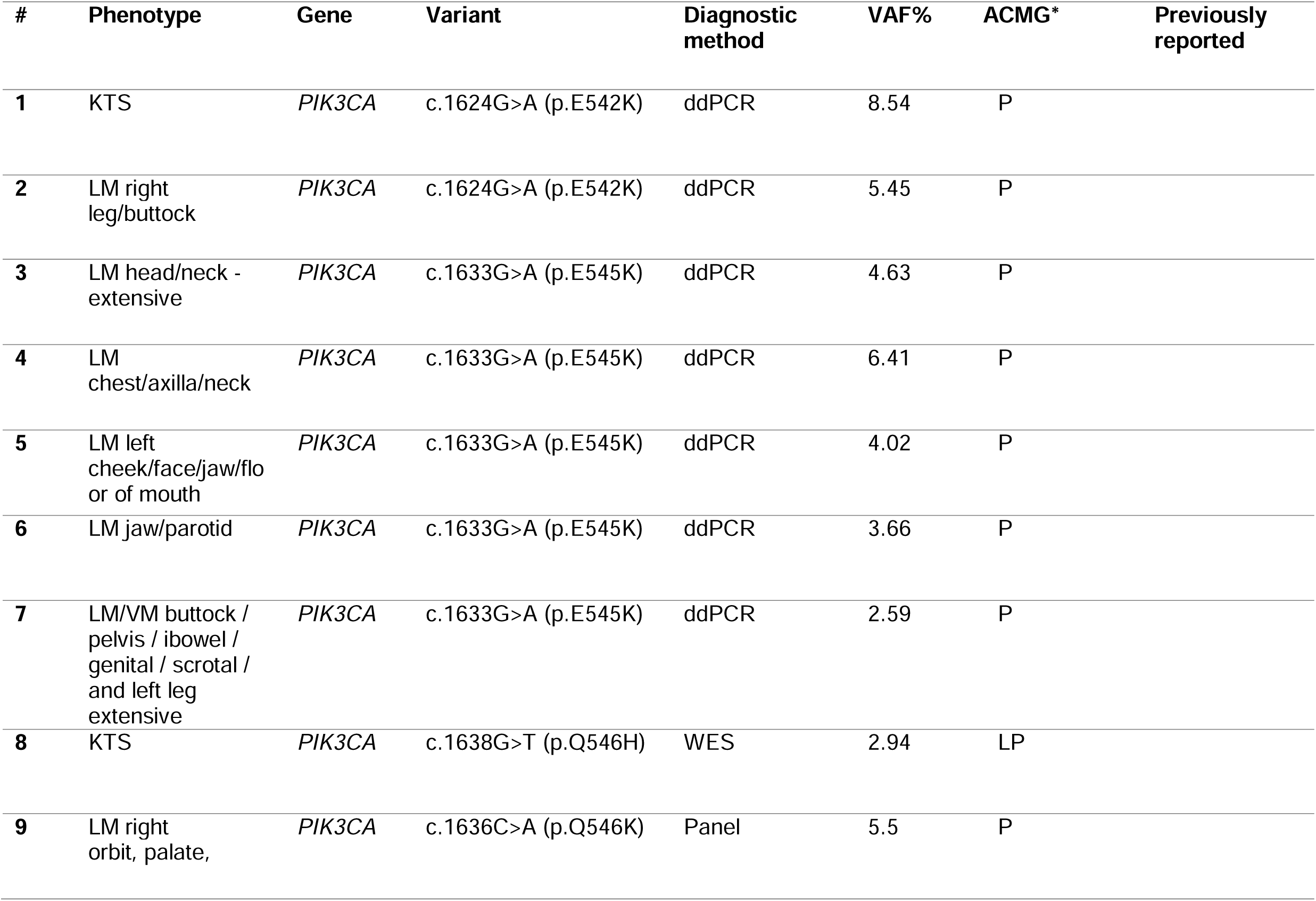
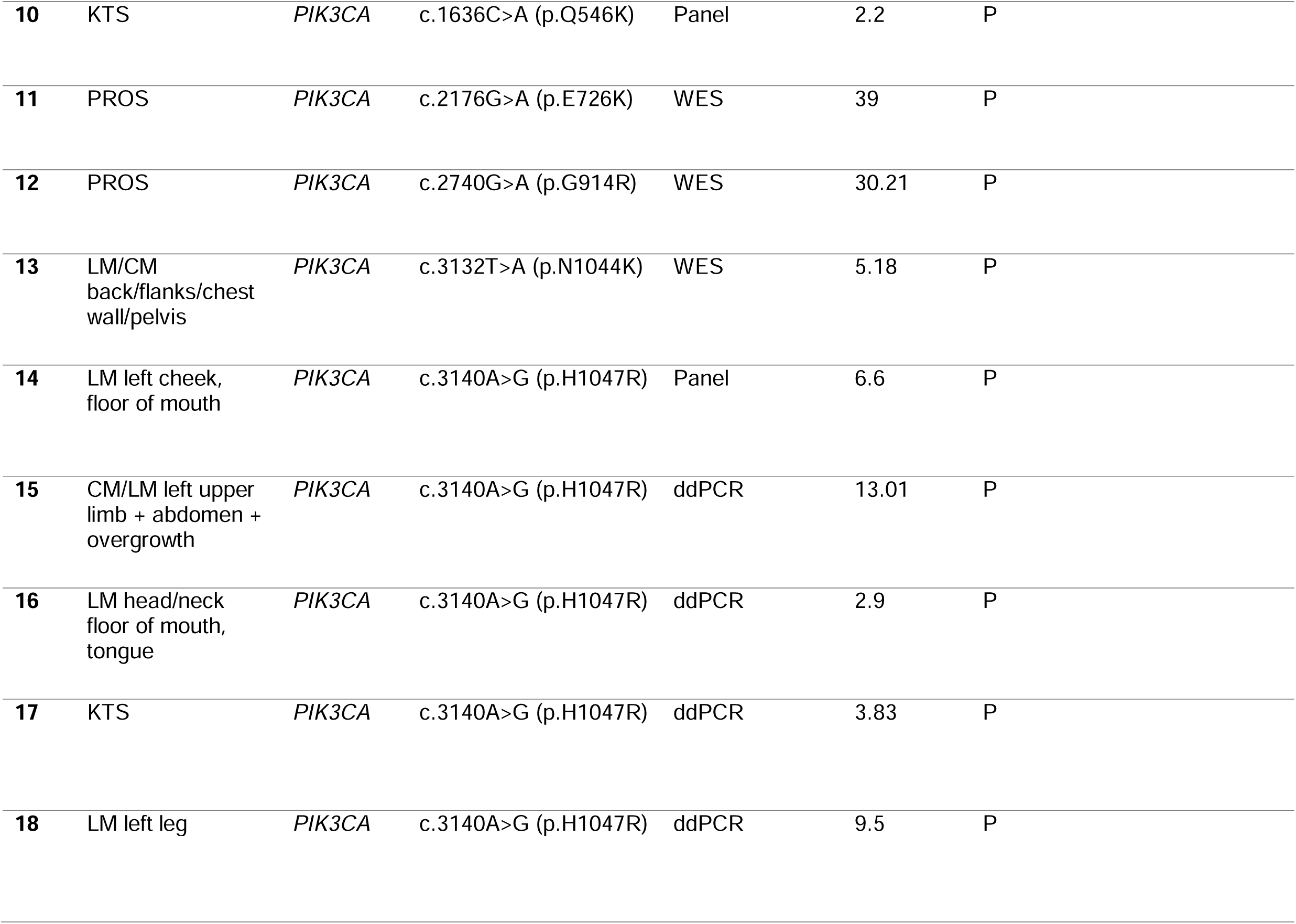
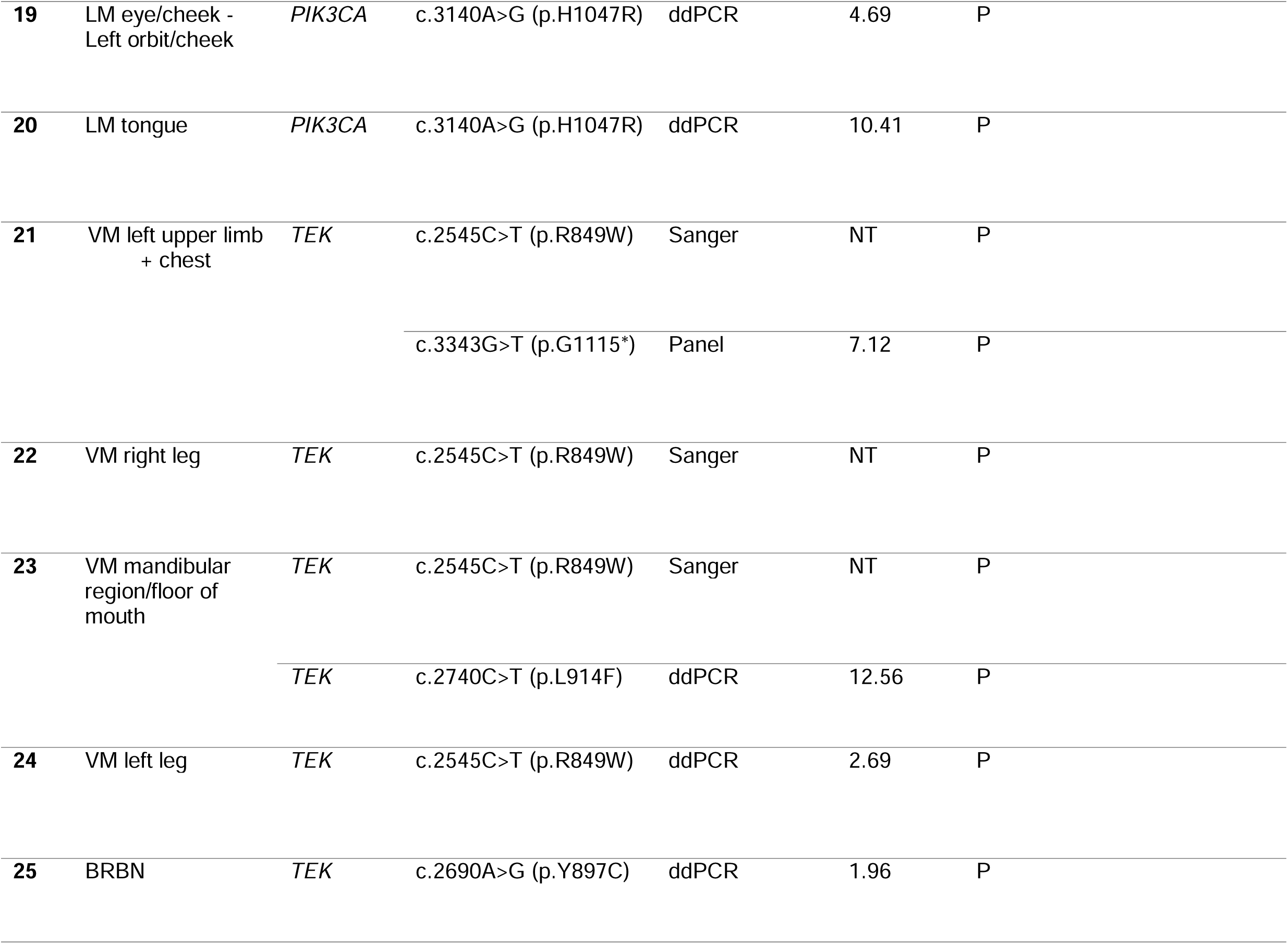
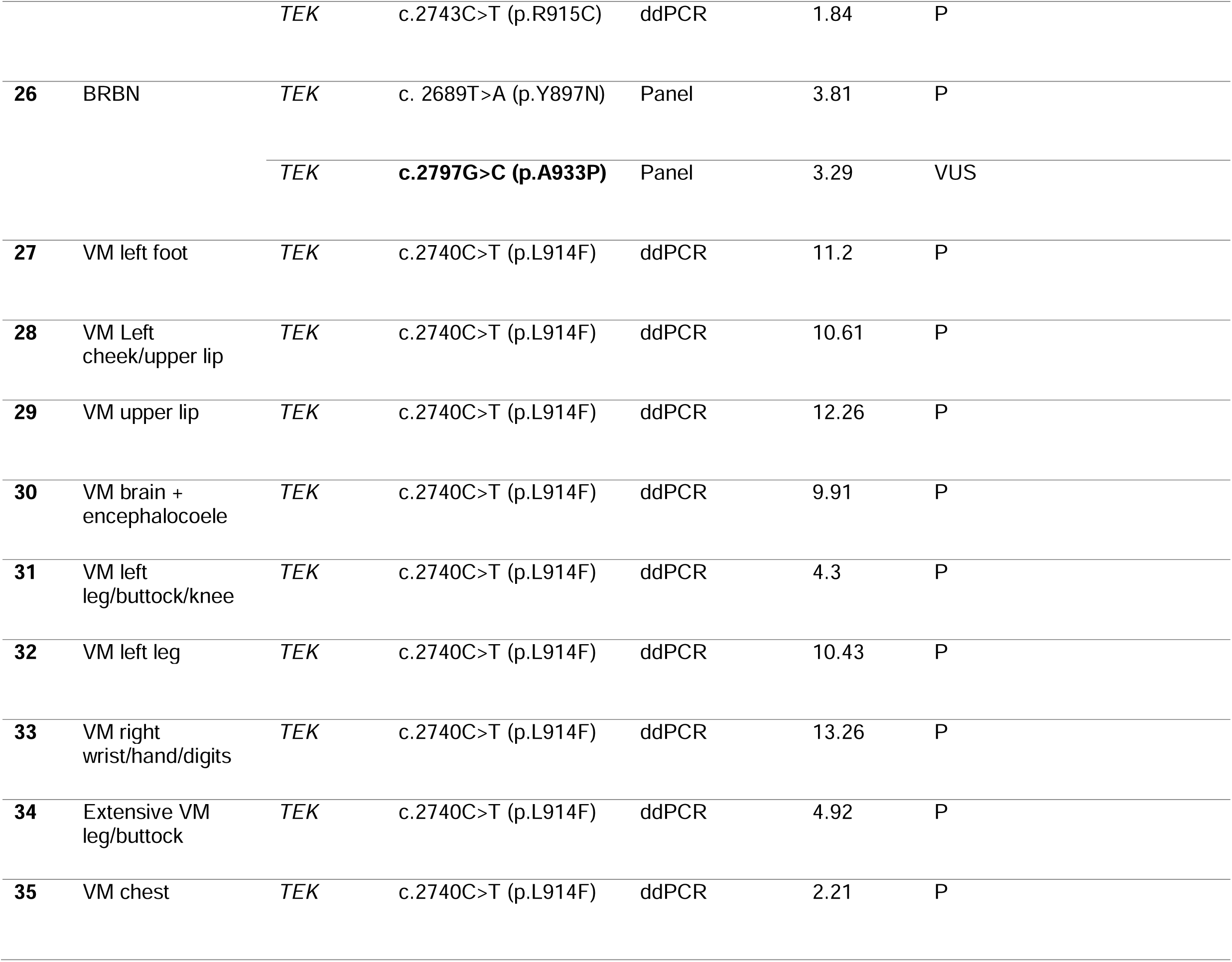
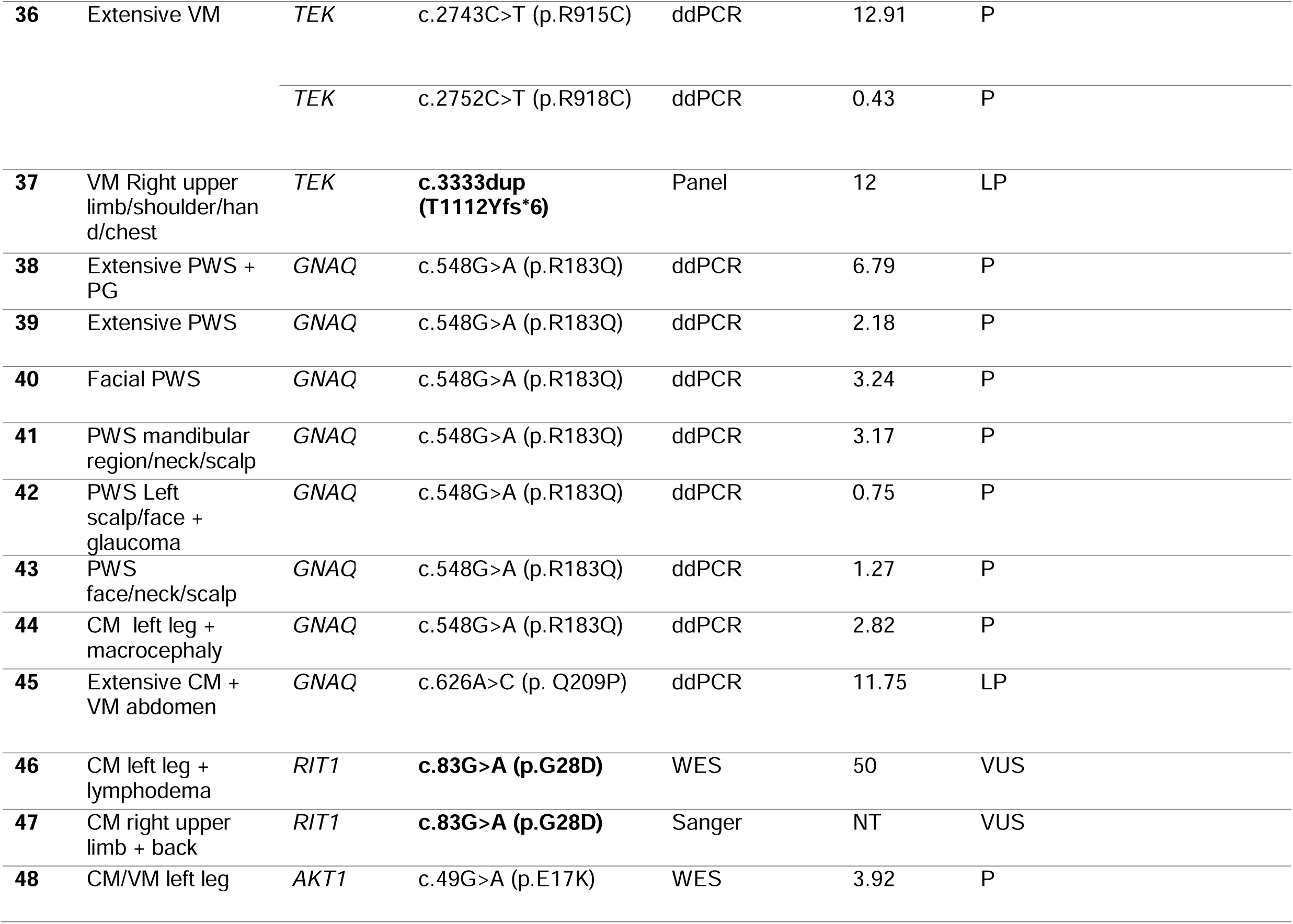
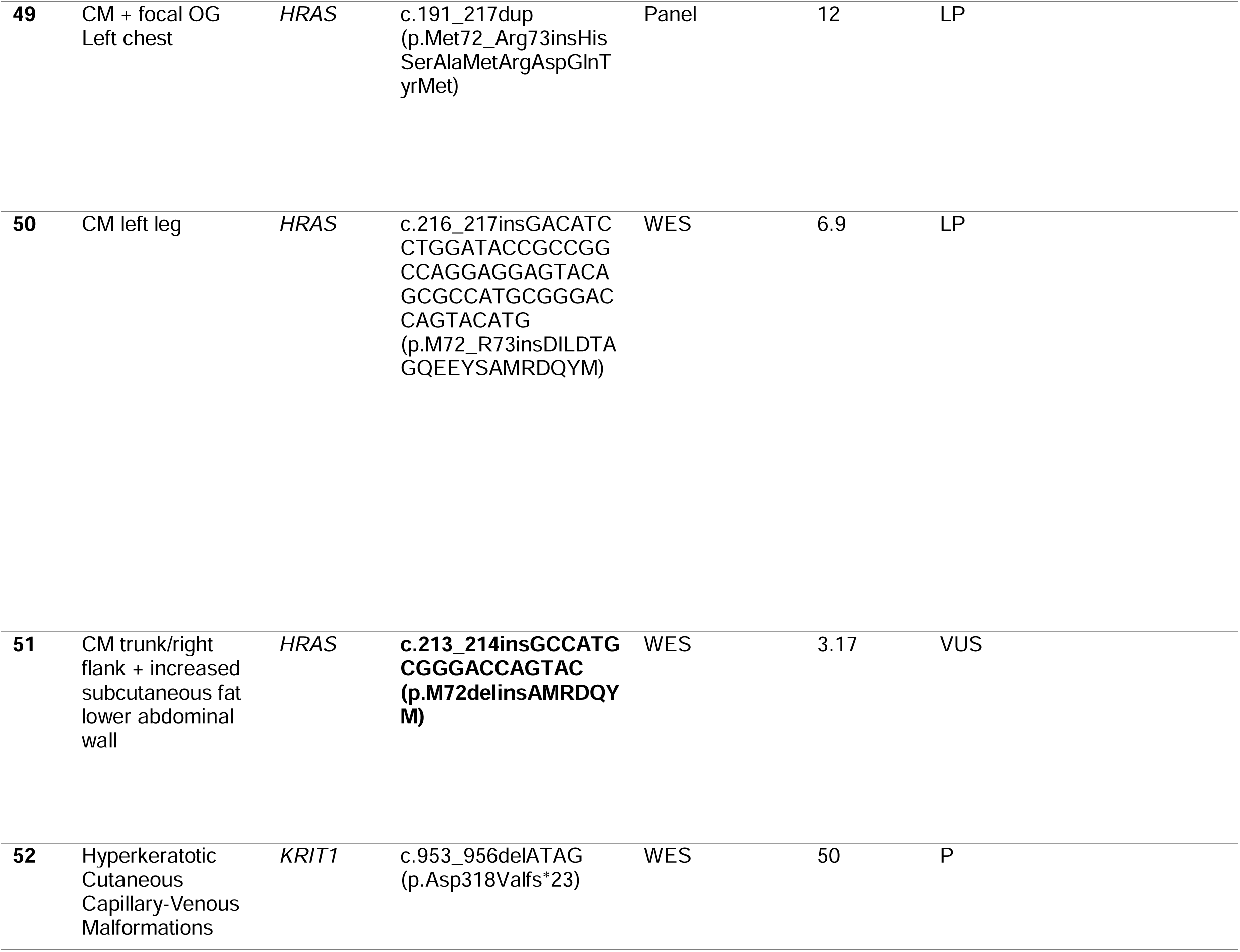
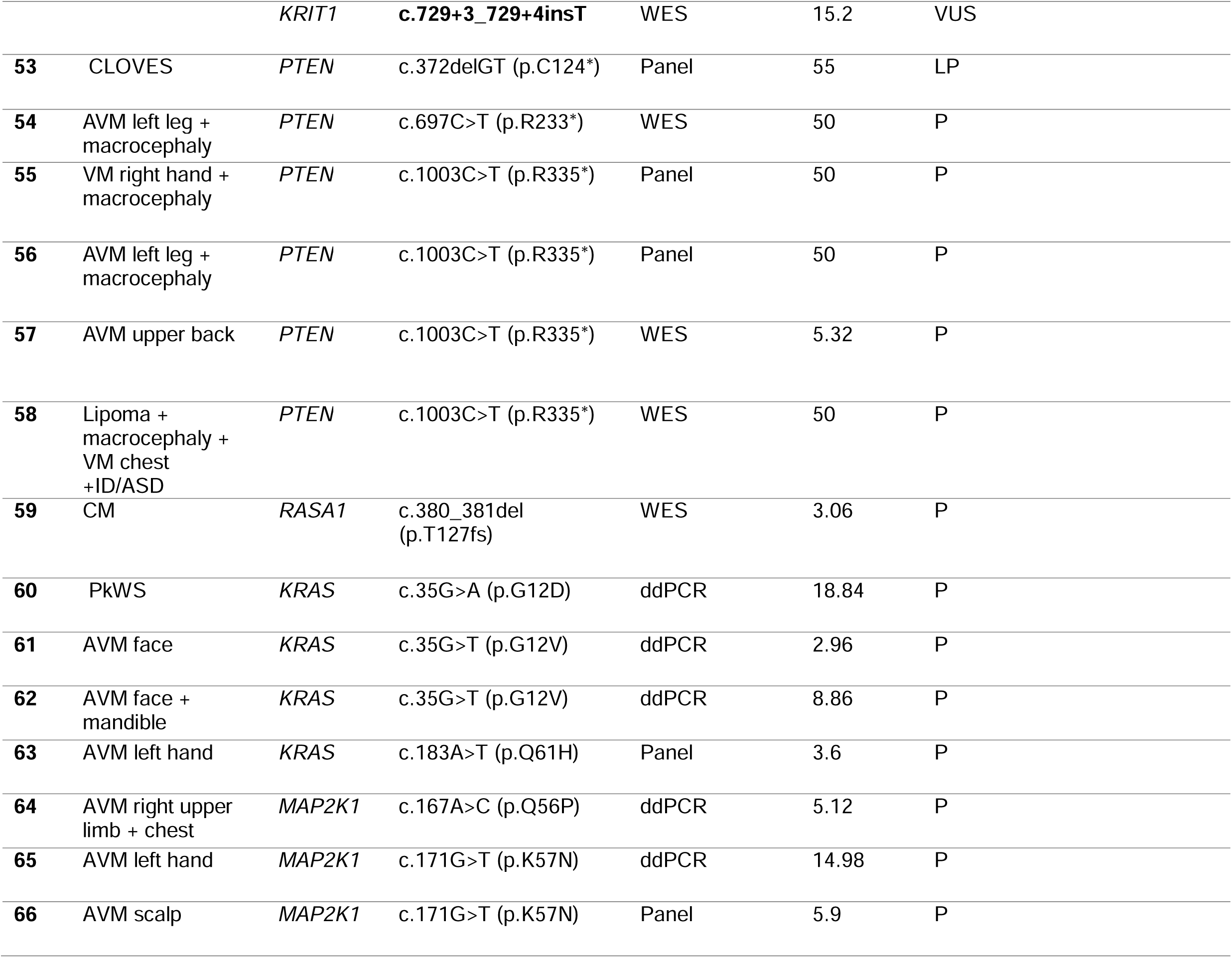
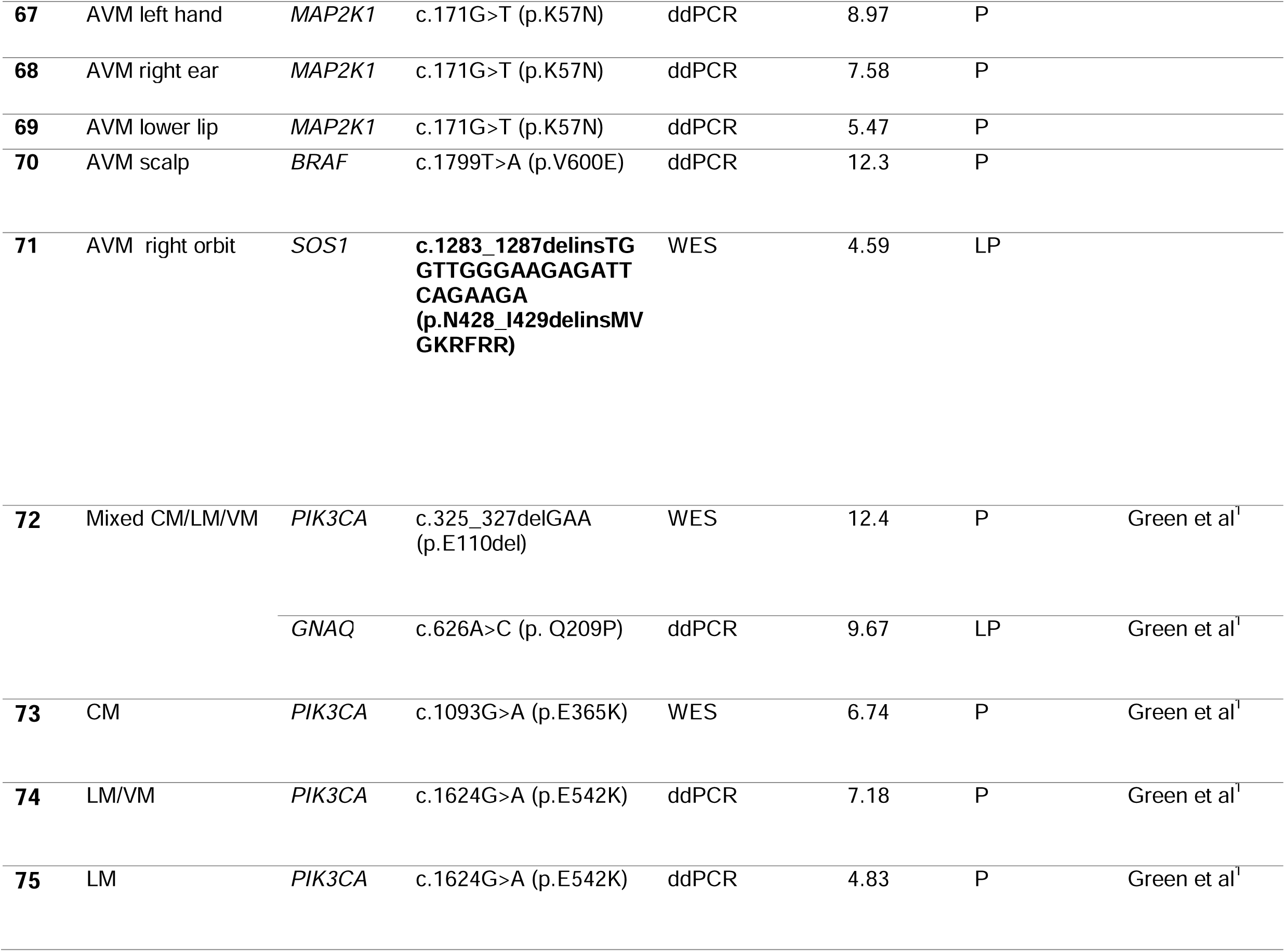
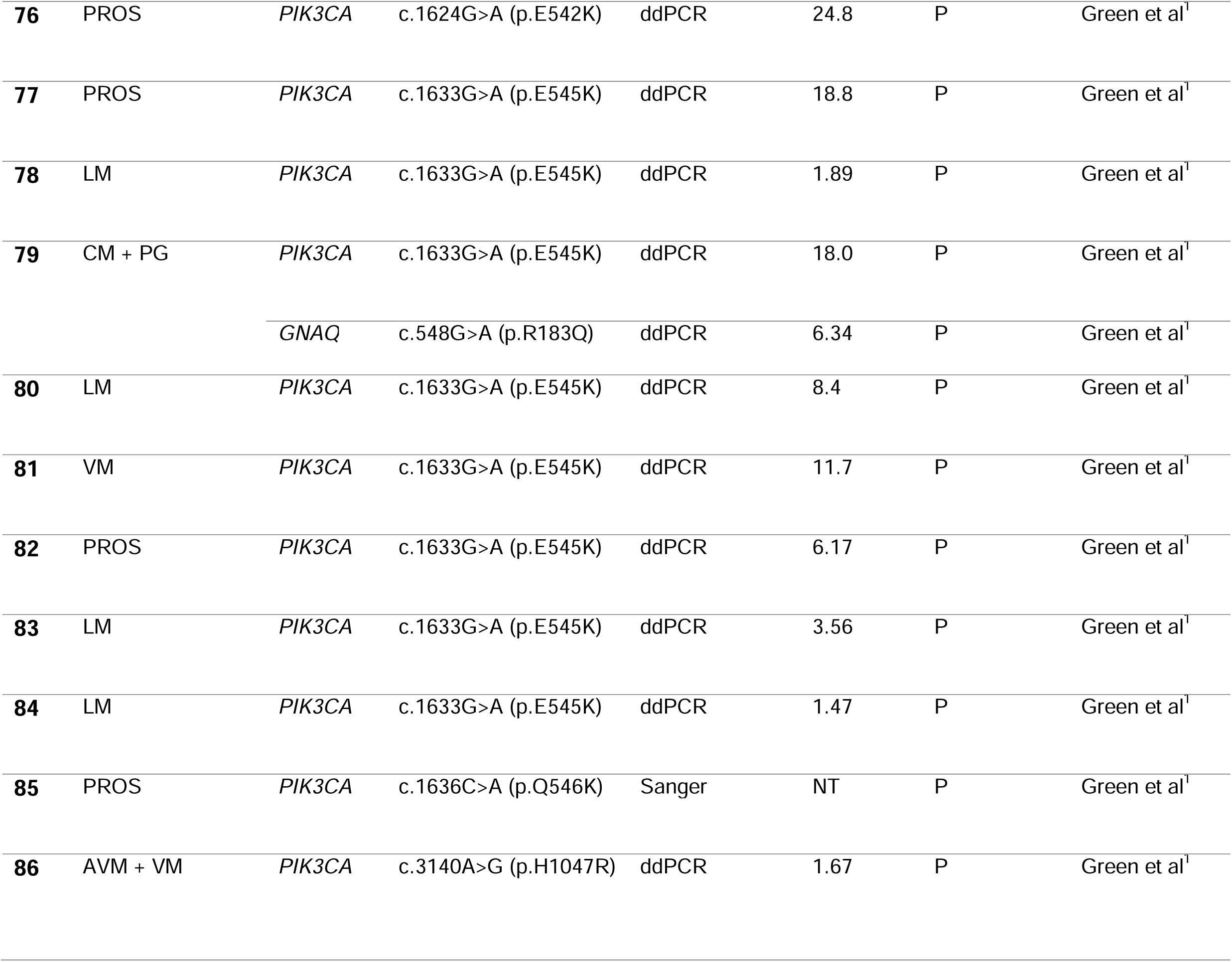
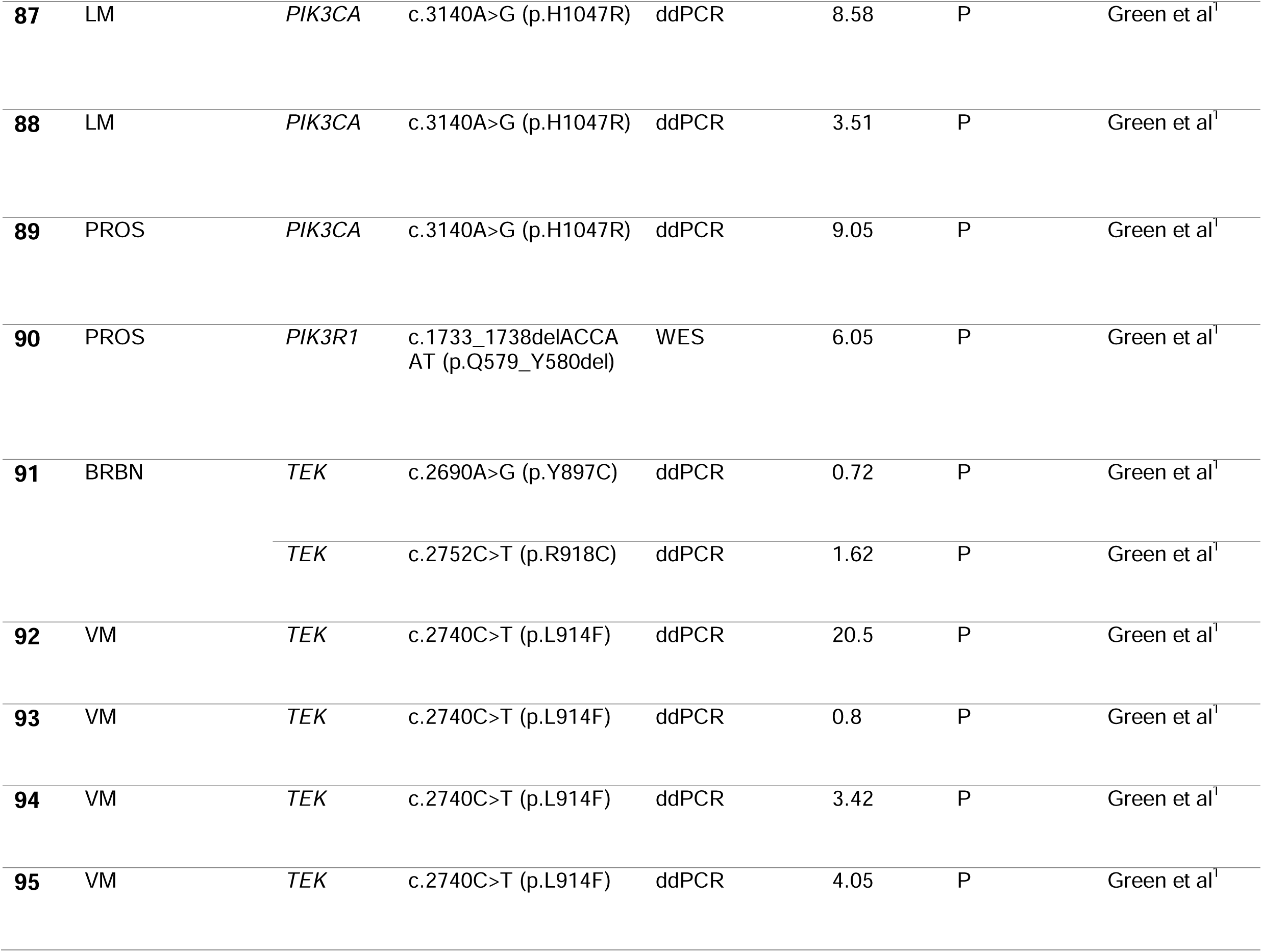
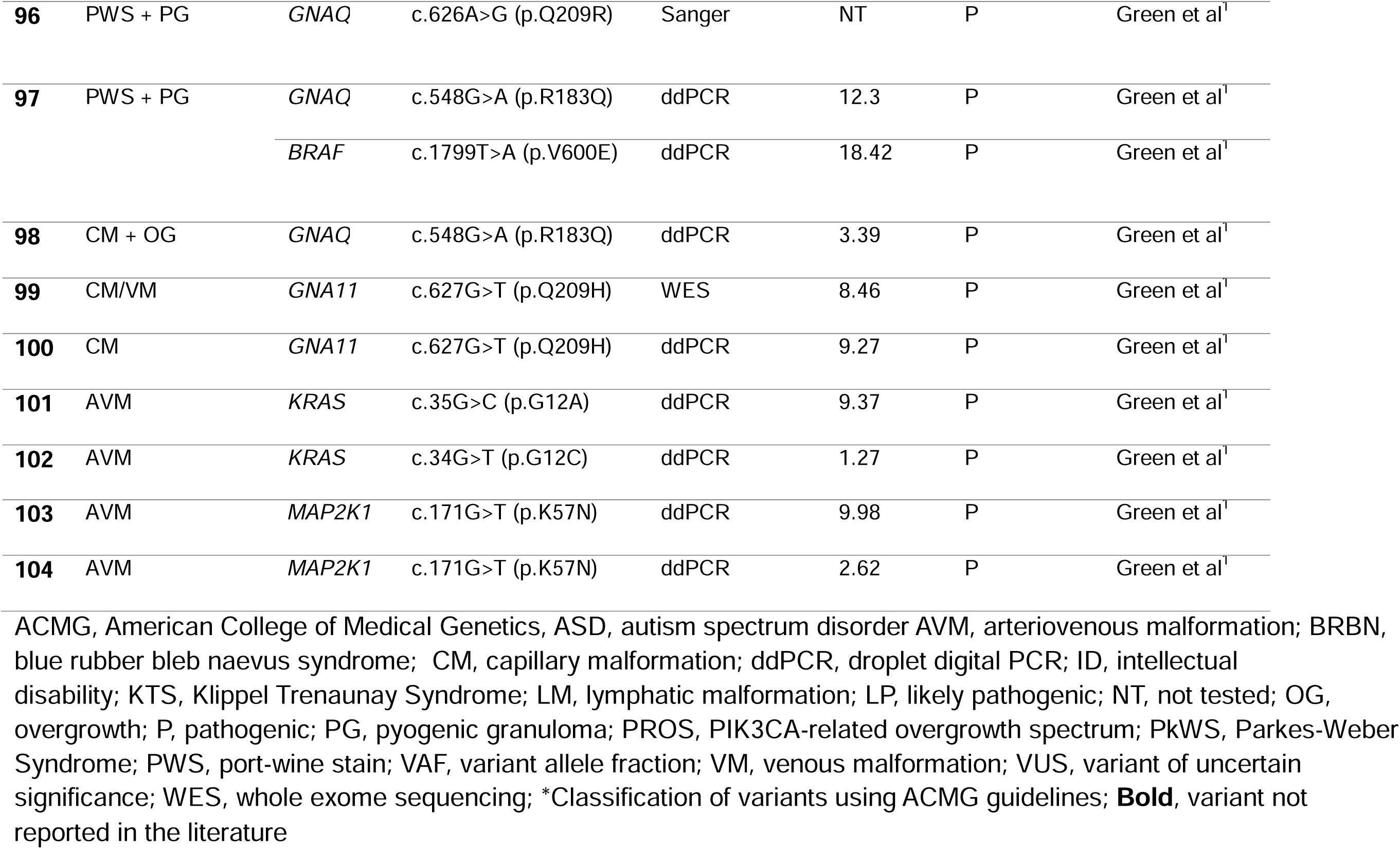
Genetic findings in 104 patients from our cohort with vascular anomalies or PROS.

We used droplet digital PCR, a highly sensitive approach to detect low-level mosaicism in lesional tissue, for first-line genetic testing. We designed specific ddPCR assays to detect 46 recurrent pathogenic variants frequently associated with non-syndromic vascular anomalies, PROS, PHTS or other syndromic forms (Supplementary Table 1). We then performed high depth sequencing using three clinical gene panels (see Supplementary Table 2 for lists of genes interrogated) or exome sequencing (see Supplementary Table 3 for list of genes interrogated) to identify variants in genes associated with vascular anomalies or novel candidate genes already associated with cancers. Generally, panels were used routinely for patients fitting clinically recognised lesion classifications, while exome sequencing was prioritised for patients with atypical or unique presentatons and suspicion of a novel gene finding. Finally, in a subset of the remaining unsolved individuals with systemic presentations or known family history of disease, we performed targeted Sanger sequencing of mutational hotspot regions (see Supplementary Table 4 for gene regions) to detect pathogenic variants beyond the highly recurrent variants already tested by ddPCR.

We detected pathogenic variants in 71/105 (∼67%) individuals by ddPCR (n=45), gene panel (n=13), exome (n=15) or Sanger (n=4), with 6 individuals having double-hit variants in *TEK* (n=5; 3 with a VM and 2 with blue rubber bleb naevus syndrome (BRBN)) or *KRIT1* (n=1, with hyperkeratotic cutaneous capillary-venous malformations and family history of cerebral cavernous malformation (CCM)) detected using multiple methods (Table 1).

Slow-flow anomalies (either lymphatic malformations (LMs), venous malformations (VMs), capillary malformations (CMs)) or PROS were present in 76/105 (∼72%) of the individuals who underwent testing. We identified somatic variants in 53/76 (∼70%) of these individuals within the genes *PIK3CA*, *TEK*, *GNAQ*, *AKT1*, *HRAS*, *KRIT1*, *PTEN* and *RASA1* with VAFs ranging from 0.75% to 39% (Table 1, Figure 2). Three individuals (1 VM, 2 BRBN) had 2 somatic variants each identified in *TEK*. We identified germline variants in 6/76 (∼8%) cases within *RIT1* (n=2), *KRIT1* (n=1), or *TEK* (n=3). Monogenic somatic second-hit variants were discovered alongside germline variants in three of these individuals, in *KRIT1* (n=1) and *TEK* (n=2).

**Figure 2.**
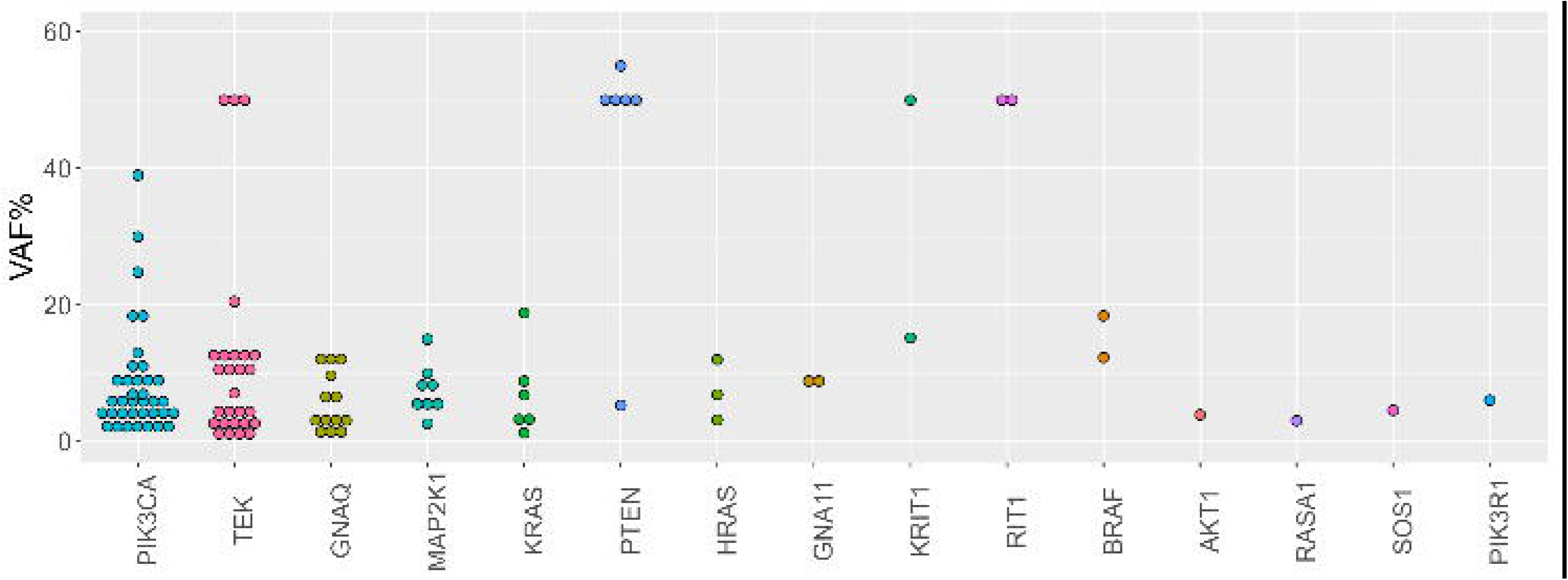
**Variant allele fraction distribution**. Variant allele fraction of somatic and germline variants detected in individuals in our cohort with vascular anomalies.

Fast-flow arteriovenous lesions were present in 15/105 (∼14%) of the individuals who underwent testing. We detected somatic variants in the genes *KRAS* (n=3), *MAP2K1* (n=6), *BRAF* (n=1), or *SOS1* (n=1) in 11/15 (∼73%) of them (Figure 3) with VAFs ranging between 2.96% to 14.98% (Table 1, Figure 2). No fast-flow cases had a germline variant.

**Figure 3.**
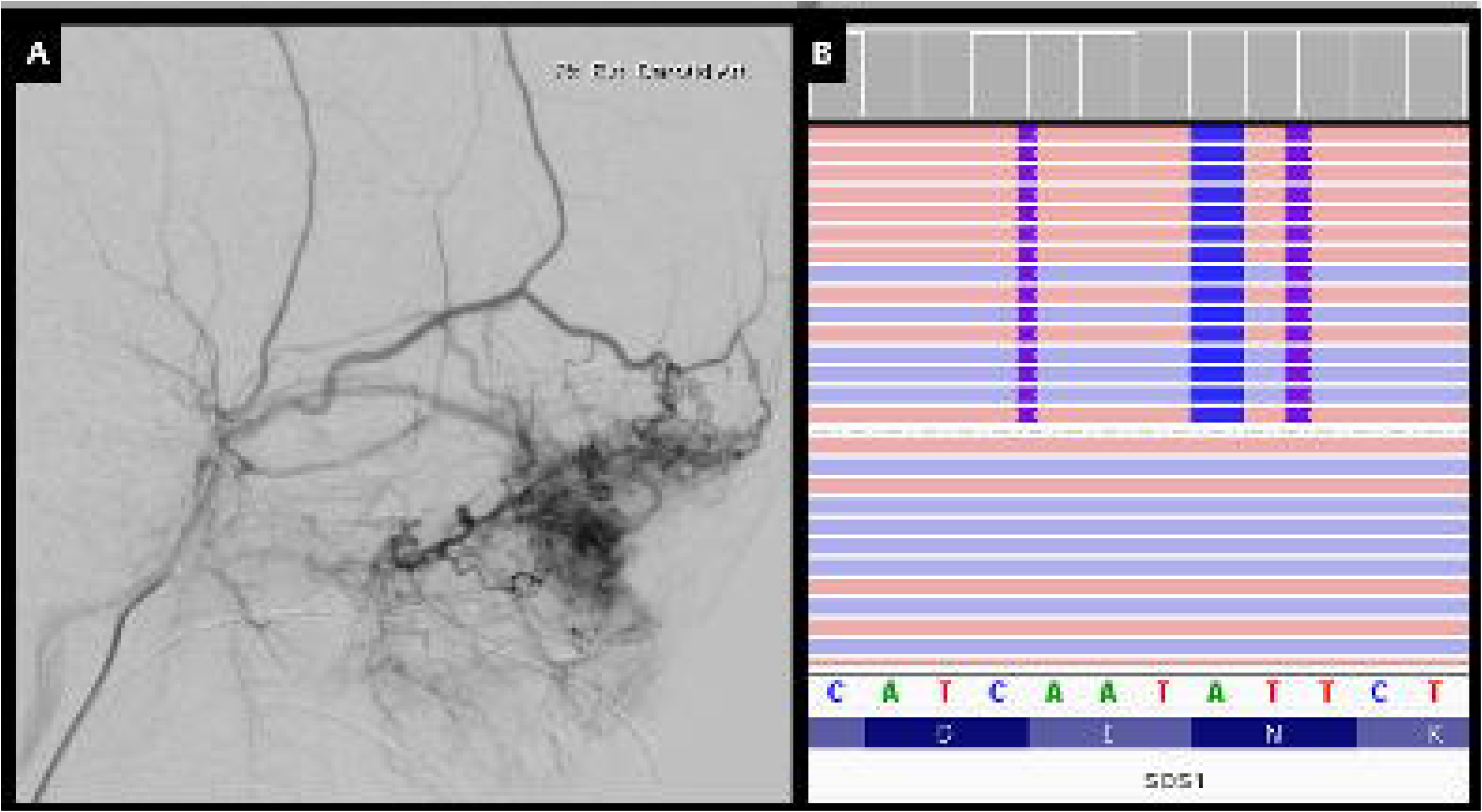
**Imaging and molecular testing for Individual 71**. Individual 71 with arteriovenous malformation of right orbital region. Angiography of external carotid artery (Panel A). Integrative Genomics Viewer screenshot showing SOS1 c.1283_1287delinsTGGTTGGGAAGAGATTCAGAAGA variant in Individual 71 (Panel B).

Mixed fast-flow and slow-flow lesions were present in 9/105 (∼9%) individuals who underwent testing. We detected somatic variants in 7/9 (∼78%) with mixed lesions. Six individuals were found to carry germline (n=5) or somatic (n=1) *PTEN* variants and subsequently received diagnoses of PHTS. One individual had an atypical mix of slow-flow (CM) and fast-flow (AVM) lesions initially suspected to be Klippel-Trenaunay (KTS) then revised to Parkes-Weber syndrome following the detection of a recurrent somatic *KRAS* variant (Table 1, Figure 1).

The cohort also included 5/105 (∼5%) individuals with a vascular tumour. Genetic testing for all five was negative.

## Discussion

Overall, when combining molecular findings from our previously reported study (n=33)^1^ and this study (n=71), the genetic diagnostic yield for our cohort was 104/138 (75%). The proportion of individuals with slow-flow lesions (81/104, ∼78%) and fast-flow lesions (14/19, ∼79%) with a molecular diagnosis was similar. Of individuals with a mix of slow-flow and fast-flow lesions, 7/9 (∼78%) had a pathogenic variant detected in *KRAS* (1/9) or *PTEN* (1/9 somatic; 5/9 germline). Surprisingly, 3/6 (50%) individuals with a *PTEN* variant did not have a prior clinical diagnosis of PHTS, suggesting this gene should be considered in genetically unsolved individuals with mixed lesions. A pathogenic variant was not found in any of the five individuals with vascular tumours.

Pathogenic somatic mosaic variants were identified in 96/138 (68%) individuals in our cohort, most at low VAFs with a mean of 7.59% [range 0.43 – 39%] (Table 1, Figure 2). Of 138 individuals tested, recurrent pathogenic variants were detected in 71 (51%) specimens on ddPCR. Variants were most frequently identified in *PIK3CA* (n=28), *TEK* (n=18), *GNAQ* (n=12), *KRAS* (n=5), or *MAP2K1* (n=7) (Figure 2). In a further 33 individuals, pathogenic variants were identified by NGS or Sanger sequencing.

Only 11/138 (8%) individuals had germline variants. These were in *TEK, PTEN, RIT1,* and *KRIT1*. In two of these individuals, Sanger sequencing was used to segregate the variant in a parent or sibling (Supplementary Figure 1). Sanger sequencing also detected two somatic variants in *PIK3CA* and *GNAQ* occurring at high variant allele fraction (Table 1). In 10/138 (7%) of individuals, multiple pathogenic variants were detected in *BRAF*, *GNAQ*, *KRIT1*, *PIK3CA* or *TEK*. These individuals carried either a germline and a somatic variant in the same gene (n=3), or two different somatic variants within a single (n=4) or multiple VA-associated (n=3) genes. Both mechanisms have been previously described in individuals with VAs, particularly in association with _CCM._17,18

The identification of a somatic *SOS1* variant in an individual with an AVM of the eyelid confirms a new gene discovery (Figure 3). Somatic *SOS1* variants associated with AVMs were also reported in abstracts presented at the 2022 and 2024 International Society for the Study of Vascular Anomalies World Congresses.^19,20^ However, these studies are yet to be published in a peer-reviewed journal. These abstracts also report AVM-linked *SOS1* variants in the critical pleckstrin homology (PH) functional domain, close to the indel variant identified in the individual in our cohort.^19^ This region may therefore represent a new mutational hotspot for AVMs. Like other AVM-associated genes (e.g., *BRAF*, *KRAS*, *MAP2K1, RIT1*), *SOS1* is a component of the Ras/Raf/MAPK pathway, and pathogenic germline variants in *SOS1* are well-described in RASopathies, including in up to 17% of individuals with Noonan syndrome.^21^ There are potential clinical implications because lymphatic malformations in individuals with SOS1-related Noonan syndrome have been shown to be responsive to treatment with MEK inhibitors.^22^

Our overall diagnostic yield (75%) equals or exceeds that of other similarly sized cohorts.^10,11,23,24^ Most of the pathogenic variants reported in these studies are highly recurrent.^8,21–23^ Over a quarter of individuals in our cohort had one of three recurrent somatic pathogenic variants (*TEK* p.L914F, *GNAQ* p.R183Q, or *PIK3CA* p.H1047R), consistent with other VA cohorts where these variants were identified in 11-33% of individuals.^10,23^ The high proportion of recurrent variants favours tiered diagnostic testing with sensitive, low-cost and efficient ddPCR assays as a first-line approach. Meanwhile, recent findings from Pakhathirathien *et al* (2025) reported a diagnostic yield of 80.8%, detecting variants in 21/26 individuals using high depth panel sequencing of 129 genes.^11^ Of the 23 P/LP variants reported across these 21 individuals, 17 (∼81%) would have been detected on our ddPCR screening panel.^11^ We estimate a reagent cost for ddPCR of around $165 USD per individual in our hands. Therefore, the cost of identifying these 17 variants by ddPCR would be ∼$2,805 USD (17 tests at $165 USD per test). Our cost for running a targeted NGS panel was $420 USD per individual. To test all 17 of these individuals on the NGS panel would have cost $7,140 USD. Thus, the overall cost saving (∼60%) would be $4,335 USD if ddPCR was used.^11^

The ddPCR yield was high in our cohort with 51% (71/138) of individuals receiving a molecular diagnosis. This reflects that ddPCR is well-suited to detecting low-level mosaicism with VAFs below 1%, a threshold below many standard clinical NGS approaches (often only designed to call variants at > 5% VAF).^25^ For example, we were able to detect a *TEK* variant in one individual at 0.43% VAF given the low detection limit of ddPCR (∼0.25%).^1^

In our previously published cohort of 60 individuals with a VA we identified a pathogenic variant in 55% (33/60) individuals.^1^ Here, we increased our diagnostic yield to 75% following testing of an additional 78 individuals. This increase reflects (i) expansion of our ddPCR assays from 29 to 46 recurrent variants (Supplementary Table 1), and (ii) improved access to high-depth clinical gene panels, including for FFPE specimens (Supplementary Table 2), that are more sensitive than exome sequencing due to increased read depth.

A molecular diagnosis may lead to a precision therapy that targets a specific pathway to improve clinical outcomes. Some of the pathogenic variants we detected here are already clinically actionable as they provide targets for existing precision therapy trials of new and repurposed oncology drugs.^26^ In fact, 12 individuals with new molecular diagnoses from this study have already been enrolled in our clinical trial of the PI3K-inhibitor alpelisib or MEK-inhibitor mirdametinib (NCT05983159).^15^ As novel therapies continue to emerge, additional individuals from our cohort with different gene variants may become eligible for future precision therapy trials.

## Methods

### Individual Recruitment and Sample Processing

Children and adults with VAs including *PIK3CA*-related overgrowth syndrome (PROS) and PTEN hamartoma tumour syndrome (PHTS) were recruited through the Royal Children’s Hospital, Melbourne, the Victorian Clinical Genetics Service, Melbourne, and St Vincent’s Hospital, Melbourne. The avaergae age of the recruited individuals was 16.7 years (range, 1.7 – 72.3 years) The Human Research Ethics Committees of the Royal Children’s Hospital [Project Number: 2019.304], Melbourne and Austin Health, Melbourne [Project Number: H2007/02961] approved this study. Informed consent was obtained from the parent or legal guardian in the case of children or the participant themselves as appropriate, for participation in the study and for the publication of clinical data including photographs including the individual depicted in Figure 3. DNA was extracted from fresh frozen or formalin-fixed paraffin embedded (FFPE) lesional tissue resections or biopsies using the QIAamp DNA FFPE Tissue Kit. In cases where germline variants were suspected, blood or saliva samples were collected and extracted using the QIAamp DNA Maxi Kit (for blood) or DNA Genotek prepIT.L2P Kit (for saliva).

### Droplet Digital PCR

All lesional samples underwent first-line ddPCR screening with TaqMan SNP Genotyping Assays from Thermo Fisher Scientific Inc. Assays were selected with consideration of the individual’s phenotype from a panel of assays designed to 46 recurrent VA-associated pathogenic variants (Supplementary Table 1). Droplet generation, PCR cycling, and droplet reading were performed according to the manufacturer’s recommendations on a Bio-Rad QX200 Droplet Digital PCR system, as we recently described (Supplemental Methods).^1^

### Next Generation Sequencing (NGS)

Clinically available gene panels from Austin Health (Melbourne, Australia), Peter MacCallum Cancer Centre (Melbourne, Australia) and Genomics for Life Pty Ltd (Brisbane, Australia) were used to interrogate panels of VA- or cancer-associated genes targeting ∼1000x coverage (Supplementary Table 2).

Deep exome sequencing was performed using the Illumina NovaSeq 6000 System (Illumina) generating 150 bp paired-end reads with 400X coverage (Beijing Genomics Institute, China). Variant filtering was used to select variants located in the exonic or splice site regions of genes on a curated list of VA genes (Supplementary Table 3), with a CADD PHRED score > 20, and gnomAD v4.1.0 allele count < 5. All variants of interest identified through NGS approaches with variant allele fractions < 50% were validated using ddPCR, as described above.

### Sanger Sequencing

Sanger sequencing was performed using primers designed to specific gene variants or hotspot regions (Supplementary Table 4) for PCR amplification and sequencing with the BigDye v3.1 Terminator Cycle Sequencing Kit (Applied Biosystems) on a 3730 XL DNA Analyzer (Applied Biosystems), using our previously reported method (Supplemental Methods).^1^

## Data Availability

Genomics data is available upon reasonable request

## Acknowledgements

The authors would like to thank the individuals and their families for participating in this study and all members of the Vascular Anomalies Clinic at the Royal Children’s Hospital.

## Data Access Statement

Genomics data is available upon reasonable request.

## Author Contributions

Conceptualisation: T.K.L., T.E.G., N.J.B., M.G.d.S., A.J.P., M.S.H.; Data Curation: T.K.L., T.E.G., N.J.B., M.G.d.S., M.F.B., A.G, R.J.P., D.M., A.C, L.P, S.J.R., P.B., J.S., A.J.P., M.S.H.; Formal Analysis: T.K.L., T.E.G., D.G., M.F.B., C.T., M.S.H.; Investigation: T.K.L., T.E.G., D.G., N.J.B., M.G.d.S., M.F.B., C.T., S.M.W.M., A.J.P., M.S.H.; Resources: N.J.B., M.G.d.S.; I.E.S., S.F.B., A.J.P., M.S.H.; Software: M.F.B.; Visualisation: T.K.L., T.E.G., N.J.B.; Writing-original draft: T.K.L., M.S.H.; Writing-review and editing: T.K.L., T.E.G., N.J.B., M.G.d.S., M.F.B., R.J.P., I.E.S., A.J.P., M.S.H.

