## Supplementary Materials for "Droplet Digital PCR as a First-Line Detection Tool in the Genetic Diagnosis of Vascular Anomalies"

**Supplemental Methods**

Droplet Digital PCR

Fluorescently-labelled droplet digital PCR (ddPCR) probes were designed and ordered from ThermoFisher Scientific (Waltham, MA). The probes were mixed with ddPCR Supermix (no dUTPs) for Probe (Bio-Rad, Hercules, CA) and 10ng of DNA. The reactions were transferred to a droplet generator cartridge with 70uL of Droplet Generating Oil for Probes (Bio-Rad) and loaded into a QX200 Droplet Generator (Bio-Rad). The droplets were then carefully pipetted into a 96-well PCR plate and transferred to a Bio-Rad C1000 Touch Thermal Cycler. PCR amplification was performed using the following conditions: initial denaturation: 95°C for 10 minutes, 38 cycles with denaturation at 94°C for 30 seconds followed by annealing at 55°C for 60 seconds, then one cycle at 98°C for 10 minutes and 12°C infinite hold. For some assays annealing temperature was adjusted within a range between 55°C and 65°C to achieve optimal primer annealing, as determined by gradient PCR. Post-PCR products were read on a QX200 droplet reader (Bio-Rad) and analyzed using QuantaSoft software. Graphical outputs of fluorescence signals are generated with R studio, creating 2D plots of amplitude and cluster data for interpretation.

Next Generation Sequencing

Exome sequencing was performed with the Agilent SureSelect DNA Human All Exon V6, 96RXN kit (Agilent Technologies) and the Illumina NovaSeq 6000 System (Illumina) with 150 bp paired-end reads targeting 400X coverage. For analysis, reads were aligned to the hg38 reference genome with BWA-MEM v0.7.17-r1188, then duplicate marking and base quality score recalibration performed with the Genome Analysis Toolkit (GATK).^1,2^ Germline variant calling was performed with GATK HaplotypeCaller and somatic variant calling with GATK Mutect2 v4.6.0.0.^1^ Variants were annotated using vcfanno and ANNOVAR.^3,4^

Variant filtering included only variants located in the exonic or splicing region of genes on a curated list of VA-causing genes, with a CADD PHRED score > 20, and gnomAD v.4.1.0 allele count < 5. All variants of interest identified through NGS approaches with variant allele fractions <50% were validated with ddPCR. Variants of interested were manually curated with Franklin (Genoox Inc). Franklin provided variant classifications based on a modified version of the American College of Medical Genetics and Genomics and Association for Molecular Pathology (ACMG/AMP) standards and guidelines for somatic mutation criteria.^5^ Variants classed as pathogenic, likely pathogenic or variant of uncertain significance were analysed for clinical relevance.

**Supplementary Figures**

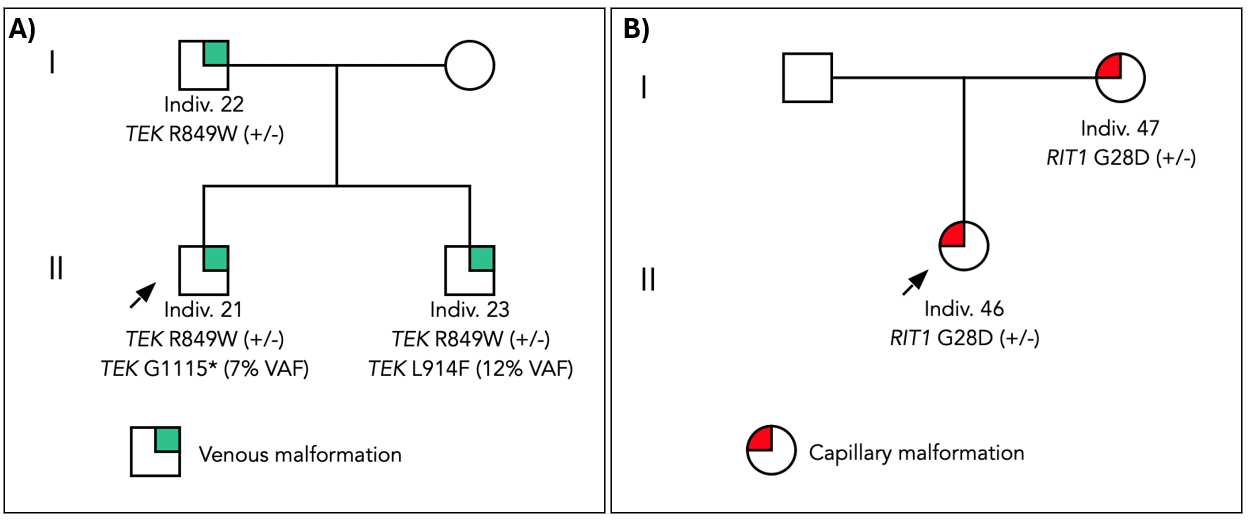

Supplementary Figure 1. A) Pedigree of familial *TEK* R849W variant; B) Pedigree of familial *RIT1* G28D variant

**Supplementary Tables**

Supplementary Table 1. Assays included in ddPCR panel of recurrent VA-linked variants

| Gene | Variant / Region |
| --- | --- |
| *PIK3CA* | c.325_327delGAA (p.E110del) |
|  | c.1093G>A (p.E365K) |
|  | c.1258T>C (p.C420R) |
|  | c.1624G>A (p.E542K) |
|  | c.1633G>A (p.E545K) |
|  | c.1636C>A (p.Q546K) |
|  | c.1638G>T (p.Q546H) |
|  | c.2176G>A (p.E726K) |
|  | c.2740G>A (p.G914R) |
|  | c.3132T>A (p.N1044K) |
|  | c.3140A>G (p.H1047R) |
| *PIK3R1* | c.1690A>G (p.N564D) |
|  | c.1699A>G (p. K567E) |
|  | c.1733_1738delACCAAT (p.Q579_Y580del) |
| *TEK* | c.2545C>T (p.R849W) |
|  | c.2689T>A (p.Y897N) |
|  | c.2690A>G (p.Y897C) |
|  | c.2740C>T (p.L914F) |
|  | c.2743C>T (p.R915C) |
|  | c.2752C>T (p.R918C) |
|  | c.2752C>A (p.R918S) |
|  | c.2797G>C (p.A933P) |
|  | c.3343G>T (p.G1115*) |
| *GNAQ* | c.110G>A (p.R37H) |
|  | c.548G>A (p.R183Q) |
|  | c.626A>C (p. Q209P) |
|  | c.626A>T (p.G209L) |
| *GNA11* | c.546_547delinsTT (p.R183C) |
|  | c.627G>T (p.Q209H) |
|  | c.626A>T (p.Q209L) |
| *GNA14* | c.614A>T (p.Q205L) |
| *AKT1* | c.49G>A (p.E17K) |
| *MAP3K3* | c.1323C>G (p.Ile441Met) |
| *PDGFRB* | c.1685A>G (p.Y562C) |
| *MET* | c.3029C>T (p.T1010I) |
| *PTEN* | c.1003C>T (p.R335*) |
| *BRAF* | c.1799T>A (p.V600E) |
| *KRAS* | c.34G>T (p.G12C) |
|  | c.35G>A (p.G12D) |
|  | c.35G>C (p.G12A) |
|  | c.35G>T (p.G12V) |
|  | c.183A>T (p.Q61H) |
| *MAP2K1* | c.167A>C (p.Q56P) |
|  | c.171G>T (p.K57N) |
| *HRAS* | c.213_214insGCCATGCGGGACCAGTAC (p.M72delinsAMRDQYM) |
| *NRAS* | c.182A>G (p.Q61R) |

Supplementary Table 2.

| Austin Health: Solid Tumour Panel | ACVR2A, AIM2, AKT1, AKT3, ALK, APC, ARID1A, ASTE1, ATM, ATP2A1, ATR, ATRX, BAP1, BARD1, BRAF, BRCA1, BRCA2, BRIP1, CBL, CCND1, CDK12, CDK4, CDKN2A, CHEK1, CHEK2, CIC, CTNNB1, DAXX, DEPDC5, DICER1, DROSHA, EGFR, EIF1AX, ELOC, EPCAM, ERBB2, ESR1, FANCA, FANCL, FGFR1, ITD, FGFR2 FGFR3, FH, FOXL2, FUBP1, GNA11, GNAQ, GNAS, GRIN2C, H3F3A, HISTH3B, HNF1A, HRAS, IDH1, IDH2, IL6ST, KBTBD4, KDM6A, KEAP1, KIT, KLHL22, KMT2C, KMT2D, KRAS, LZTR1, MAP2K1, MAP3K3, MARCKS, MDM2, MEN1, MET, MITF, MLH1, MRE11, MSH2, MSH6, MTOR, MYB, MYC, MYCN, NBN, NF1, NF2, NPRL2, NPRL3, NRAS, PALB2, PBRM1, PDGFRA, PIK3CA, PIK3R1, PMS2, POLE, PPFIA4, PRKAR1A, PRKCA, PRKD1, PTEN, PTHLH, PTPN11, RAD51A, RAD51B, RAD51C, RAD54L, RAF1, RB1, RET, RHEB, RIT1, RNF43, ROS1, SDHA, SDHB, SDHC, SDHD, SF3B1, SLC35A2, SMAD4, SMARCA4, SOS1, STK11, TAF1B, TEK, TERT, TFEB, TGFBR2, TP53, TSC1, TSC2, Fusions: ALK, RET, ROS1, CCDC6, EML4, NCOA4, |
| --- | --- |
| Genomics For Life: Vascular Malformations and Overgrowth Panel | ACVRL1, AKT1, AKT3, BRAF, CCM2, CCND2, CDKN1C, ELMO2, ENG, EPHB4, GDF2, GJA4, GLMN, GNA11, GNA14, GNAQ, HRAS, KRAS, KRIT1, MAP2K1, MAP3K3, MTOR, NRAS, PDGFRB, PIK3CA, PIK3R1, PIK3R2, PTEN, RASA1, SMAD4, STAMBP, TEK |
| Peter MacCallum Cancer Centre: PeterMac Oncomine Precision Assay | AKT1, AKT2, AKT3, ALK, AR, ARAF, BRAF, CDK4, CDKN2A, CHEK2, CTNNB1, EGFR, ERBB2, ERBB3, ERBB4, ESR1, FGFR1,  FGFR2, FGFR3, FGFR4, FLT3, FOXL2, GNA11, GNAQ, GNAS, H3-3A, H3C2, H3C3, HRAS, IDH1, IDH2, KIT, KRAS, MAP2K1, MAP2K2, MET, MTOR, NRAS,  NTRK1, NTRK2, NTRK3, PDGFRA, PIK3CA, POLE, PTEN, RAF1, RET, ROS1, SMO, TERT promoter, TP53*; copy number alterations in ALK, AR, CD274,  CDKN2A, EGFR, ERBB2, ERBB3, FGFR1, FGFR2, FGFR3, KRAS, MET, PIK3CA, PTEN, 1p/19q co-deletion; gene fusions involving ALK, BRAF, ESR1,  FGFR1, FGFR2, FGFR3, MET, NRG1, NTRK1, NTRK2, NTRK3, NUTM1, RET, ROS1, RSPO2, RSPO3; and splice variants in AR, EGFR and MET. |

### Supplementary Table 3. List of VA-linked genes used to filter variants in WES analysis

| *ACVRL1* | *CCM2* | *FLT4* | *GNAQ* | *KRAS* | *PDGFRB* | *SMAD4* |
| --- | --- | --- | --- | --- | --- | --- |
| *ADAMTS3* | *CCND2* | *FOXC2* | *GNB2* | *KRIT1* | *PIEZO1* | *SOS1* |
| *AGGF1* | *CDKN1C* | *GATA2* | *HGF* | *LMPH1B* | *PIK3CA* | *SOX18* |
| *AKT1* | *CDH11* | *GDF2* | *HRAS* | *MAP2K1* | *PIK3R1* | *STAMBP* |
| *AKT2* | *CELSR1* | *GJA1* | *IDH1* | *MAP3K3* | *PIK3R2* | *TEK* |
| *AKT3* | *DDX24* | *GJA4* | *IDH2* | *MET* | *PLCG1* | *TNFRSF11A* |
| *ANGPT2* | *ELMO2* | *GJC2* | *IKBKG* | *MTOR* | *PTPRB* | *TREM2* |
| *BAD* | *ENG* | *GLMN* | *IRS2* | *NF1* | *PTEN* | *VEGFC* |
| *BRAF* | *EPHB4* | *GNA11* | *KDR* | *NRAS* | *PTPN14* | *RIT1* |
| *CCBE1* | *FAT4* | *GNA14* | *KIF11* | *PDCD10* | *RASA1* | *FOXO1* |

Supplementary Table 4. PCR Primers used for Sanger sequencing

| Region | Primer Sequence 5’–3’ |
| --- | --- |
| *GNAQ* exon 5 | Forward:  GCTTAGAGTTCGAGTCCCCA |
|  | Reverse:  TGGGGTCCATCATATTCTGG |
| *RIT1* exon 2 | Forward: CCTCCTTTTCTAGGTGGCTTTTC |
|  | Reverse:  CTGGGAATCGGTGGCTGAT |
| *PIK3CA* exon 8 | Forward:  AAATATCTCATGCTTGCTTTGGT |
|  | Reverse:  GAGAGAAGGTTTGACTGCCATAA |
| *PIK3CA* exon 10 | Forward:  ATCTGGTCTTGTTGTTGGCT |
|  | Reverse:  TACCCGTATCACCAACAGCA |
| *PIK3CA* exon 21 | Forward:  TGCTCCAAACTGACCAAACTG |
|  | Reverse:  GCTATCAAACCCTGTTTGCG |
| *TEK* exon 15 | Forward:  GGATGCCAACCAGAAGACAT |
|  | Reverse:  GTTTTCTCCACACCCTCACG |
| *TEK* exon 17 | Forward:  AGGCAATTTCCACAGCACAT |
|  | Reverse:  GAGAGCTTAAGGTACCTCGCT |
